# High- versus low-dose dietary n-3 PUFA treatment produces mixed effects on DNA methylation and epigenetic fidelity in breast adipose tissue

**DOI:** 10.64898/2026.03.18.26348746

**Authors:** David E Frankhouser, Hongwei Holly Yin, Martha A Belury, John W Newman, Lisa D Yee

## Abstract

Long-chain n-3 polyunsaturated fatty acids (n-3 PUFAs) are candidate preventive agents for breast cancer. With emerging evidence of epigenetic regulation of the tumor microenvironment, tissue-level epigenetic effects may represent an important target for cancer prevention. In a randomized Phase II sub-study (high-dose 5 g/day vs low-dose 1 g/day for 12 months; n = 17; *Clinicaltrials.gov*: NCT02295059), DNA methylation (DNAm) of the breast environment was profiled by reduced-representation bisulfite sequencing (RRBS). DNAm was assessed genome-wide, at individual gene promoters, and for locus-level heterogeneity which has been linked to epigenetic dysregulation that can precede breast cancer. Both doses induced promoter DNAm changes, but their responses diverged: low-dose samples showed increased CpG variance and more differentially methylated promoters without pathway enrichment, whereas high-dose samples had reduced DNAm heterogeneity and promoter enrichment in inflammation signaling pathways. Many overlapping differentially methylated promoters changed in opposite directions between doses. The finding that high-dose n-3 PUFA affects DNAm fidelity in the breast adipose suggests a new potential mechanism for n-3 PUFA-mediated prevention of breast cancer development. Together with the dose-specific, directionally discordant DNAm responses in breast adipose, this study has important implications for both advancing n-3 PUFA for breast cancer prevention and dose selection in future n-3 PUFA supplementation trials.

## INTRODUCTION

Increasing evidence indicates a key role for breast adipose tissue in modulating the risk of mammary carcinogenesis. As a major component of the mammary microenvironment, adipose tissue serves as a source of essential fatty acids, adipokines (e.g., adiponectin and leptin), proinflammatory cytokines (e.g., tumor necrosis factor alpha), fatty acid derived metabolites, and other related factors. Dysfunctional metabolic and endocrine signaling in adipose tissue may engender a cancer-prone microenvironment with aberrant cell signaling and crosstalk. For example, obesity and high adiposity are linked to chronic inflammation conducive to development of solid malignancies such as prostate, colorectal, endometrial, hepatocellular, and breast cancers.

Breast adipose fatty acid composition is influenced by dietary fat intake. Based on epidemiological studies, diets associated with increased breast cancer risk and disease progression include high fat diets as well as diets high in saturated fatty acids.^1–4^ The long chain marine omega-3 (n-3) polyunsaturated fatty acids (PUFAs), eicosapentaenoic acid (EPA) and docosahexaenoic acid (DHA), have emerged as promising candidates for breast cancer prevention based on extensive preclinical evidence including our own studies in a murine model of hormone receptor negative, human epidermal growth factor receptor 2 (HER-2) positive breast cancer.^5–9^ EPA and DHA exert anti-inflammatory effects through multiple mechanisms, including inhibition of pro-inflammatory cytokine production and promotion of macrophage-mediated clearance of apoptotic cells.^10–14^ Additionally, n-3 PUFAs can directly influence cell membrane composition, alter membrane fluidity, modulate lipid raft formation, and undergo metabolism to bioactive by-products with anti-inflammatory properties.^15,16^ Conversely, n-6 PUFAs have been linked to increased tumor growth and metastasis in rodent models of mammary tumorigenesis.^17,18^ As a dietary intervention with multiple molecular and metabolic effects, the anti-cancer mechanism in humans is likely complex, multi-factorial, and dependent on baseline genetic, epigenetic, and phenotypic state.

Recent research to identify the causes of breast cancer initiation and development has highlighted the critical role of epigenetic alterations, particularly DNA methylation (DNAm). Aberrant DNAm patterns and increased epigenetic heterogeneity in benign breast tissue have been identified as an early event that even precedes breast cancer development.^19,20^ These epigenetic alterations frequently affect genes involved in cellular differentiation, proliferation, and inflammatory responses, suggesting a mechanistic link between epigenetic dysregulation and cancer development.^21–24^ Unique epigenetic profiles in adipose tissue have also been identified in individuals with obesity^25^ and raise the potential for aberrant DNAm patterns in breast adipose tissue as a key component of the mammary microenvironment. Notably, recent studies have demonstrated abnormal adipose tissue biology as mediated by DNAm in adipose tissue adjacent to colorectal cancer (i.e., visceral adipose tissue) and prostate cancer (i.e., periprostatic adipose).^26,27^

In our recent Phase II randomized dose response trial in breast cancer survivors of one-year treatment with n-3 PUFAs,^28^ breast adipose tissue analyses demonstrated greater increases in n-3 PUFA content and EPA-, DHA-derived oxylipins with high-dose (5 g EPA+DHA/d) relative to low-dose (1 g EPA+DHA/d) supplementation, as well as breast adipose DNAm patterns indicating downregulation of at least two metabolic pathways involved in mammary carcinogenesis.^28^ By comparing the DNAm results observed in the high-dose arm to the low-dose arm, we determined n-3 PUFA effects unique to the high-dose arm and identified complex dose-specific effects with important implications for the design of future prevention trials.

## RESULTS

### Differential effects on global DNAm after high-dose or low-dose n-3 PUFAs

This DNAm sub-study used randomly selected adipose tissue samples (n =17) from a Phase II dose-response clinical trial (*Clinicaltrials.gov:* NCT02295059) of n-3 PUFAs.^28^ The primary analysis showed that high-dose relative to low-dose n-3 PUFA treatment resulted in significantly higher breast adipose n-3 PUFA fatty acids and related fatty acid metabolites and significantly increased erythrocyte membrane and plasma n-3 PUFAs and decreased serum triglyceride levels.^28^ As with the full study cohort (n =51), the high-dose samples of this DNAm sub-study also showed significantly increased n-3 PUFA content over one year compared to the low-dose arm for both breast adipose tissue (**Fig. 1A;S1A**) and RBCs (data not shown).^28^ Serum triglyceride levels decreased from baseline to 6 and 12 months in only the high-dose arm (**Fig. 1A**). Taken together, these changes all demonstrated dose-dependence with larger changes in the high-dose relative to the low-dose arm.

**Figure 1.**
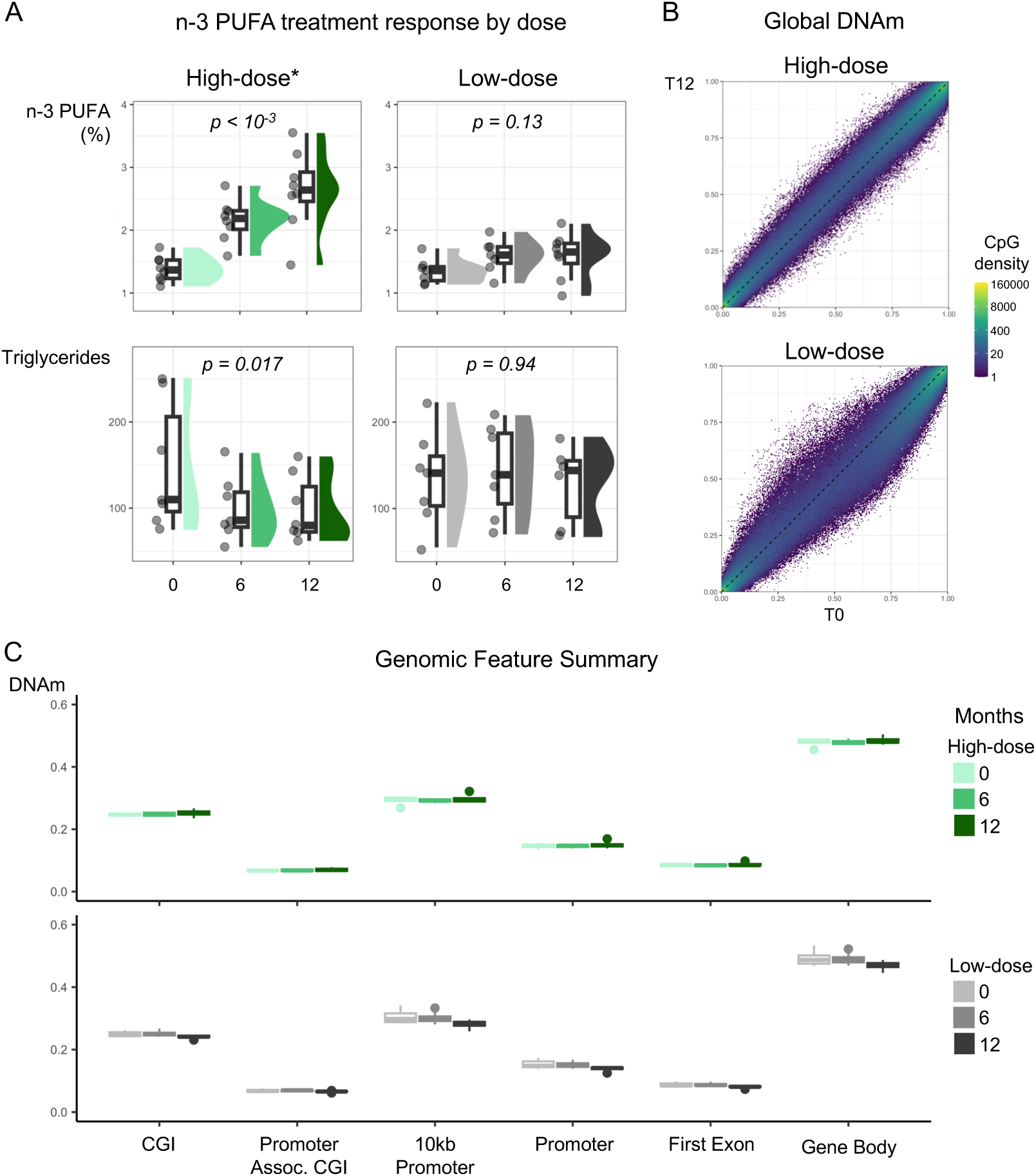
Global DNAm changes compared between treatment arm. **A)** Comparison of n-3 PUFA and triglycerides changes at 0, 6, and 12-month high- or low-dose n-3 PUFA treatment for the samples in this sub-study (n=7 high-dose, n=10 low-dose participants). **B)** Comparison of global DNAm between 0 and 12-months for both high- and low-dose treatments. Average DNAm for each CpG and all CpGs plotted using the color of each point to indicate the number of CpGs. **C)** DNAm change tested in annotated genomic features to determine whether DNAm changed between 0 and 6-months or 0 and 12-month for each dosing arm. No changes were observed in the high-dose arm; unpaired t-test showed decreases in DNAm in the low-dose 0 vs 12-month comparisons (pval < 0.05 in 10kb Promoter, Promoter, and Gene Body) that were not significant after FDR multiple test correction.

To assess the effects of n-3 PUFA supplementation on breast adipose DNAm, we utilized reduced representation bisulfite sequencing (RRBS) to analyze samples collected at 0 (baseline), 6, and 12 months after high- and low-dose n-3 PUFA treatment. For genome-wide analysis, we first compared the global change in DNAm at each cytosine-phosphate-guanine (CpG) dinucleotide with sufficient read coverage by plotting baseline relative to 12mo post-treatment DNAm values for each arm (**Fig. 1**). Fitting the resulting plots did not detect global changes in DNAm from 0 and 12 months for either dose arm (**Fig. 1B**; **Fig. S2A**). However, the low-dose arm had greater variance in DNAm after treatment, quantified both by 1) measuring the width at the 99^th^ percentile bands on each plot which showed the low-dose arm had a wider distribution of average DNAm values (**Fig. S2B**) and 2) performing an ANOVA analysis of DNAm differences which showed a significant difference (p <0.001; **Fig. S2C**). The global DNAm trajectories of each participant showed relatively small fluctuations in DNAm over the course of n-3 PUFA treatment without dose effects (**Fig. S2A**).

To determine whether n-3 PUFA treatment induced DNAm changes in specific regions of the genome, we next quantified DNAm in different annotated genomic features selected for relevance to DNAm machinery and DNAm regulation of gene expression. To compare the effects of n-3 PUFAs in these features, the average DNAm was calculated for each arm and timepoint (**Fig. 1C**). DNAm in the high-dose arm did not significantly change from baseline at 6 and 12 months in these regions. Although the low-dose arm had DNAm decreases in the promoters and first exon, none of the changes achieved significance (**Fig. 1C**). Likewise, the genomic distribution of DNAm around the transcriptional start site did not change after n-3 PUFA treatment in either arm (**Fig. S2D**).

Together, these global assessments of DNAm did not reveal significant genome-wide DNAm changes in the either arm; however, we observed increased DNAm variability in the low-dose arm after 12 months (**Fig. 1B; Fig. S2B-C**).

### Differentially methylated regions in gene promoters in breast adipose tissue vary inversely between high-dose and low-dose n-3 PUFA treatment

To investigate n-3 PUFA treatment effects on DNAm of specific genes and pathways in breast adipose tissue, we analyzed gene promoters for differentially methylated regions (DMRs) defined as promoter regions with both a statistically significant multiple-test corrected p-value and minimum DNAm change of 5%.^29^ At 12 months of study treatment, the low-dose arm had more DMRs than the high-dose arm (1680 low-dose vs 638 high-dose DMRs). For each arm, we compared DNAm between the baseline and both 6- and 12-months post-treatment (**Fig. 2A**). The direction of DNAm changes with respect to the pre-treatment timepoint classified DMRs as either hypo- or hyper-methylated. Most DMRs (n =1540) were unique to each dosing arm, with 389 DMRs shared by both arms (**Fig. 2B; Fig. S3A**); however, over 74% (288/389) of shared DMRs had DNAm that changed in opposite directions (**Fig. 2B**). Additionally, the DMRs trended in different directions between dosing arms at 12 months: the high-dose arm DMRs were enriched for hypermethylation, whereas the low-dose arm DMRs were enriched for hypomethylation (hypergeometric pval < 0.05). Taken together, DMR analysis of breast adipose DNAm revealed a complex, non-linear, and directionally discordant response to n-3 PUFA dose.

**Figure 2.**
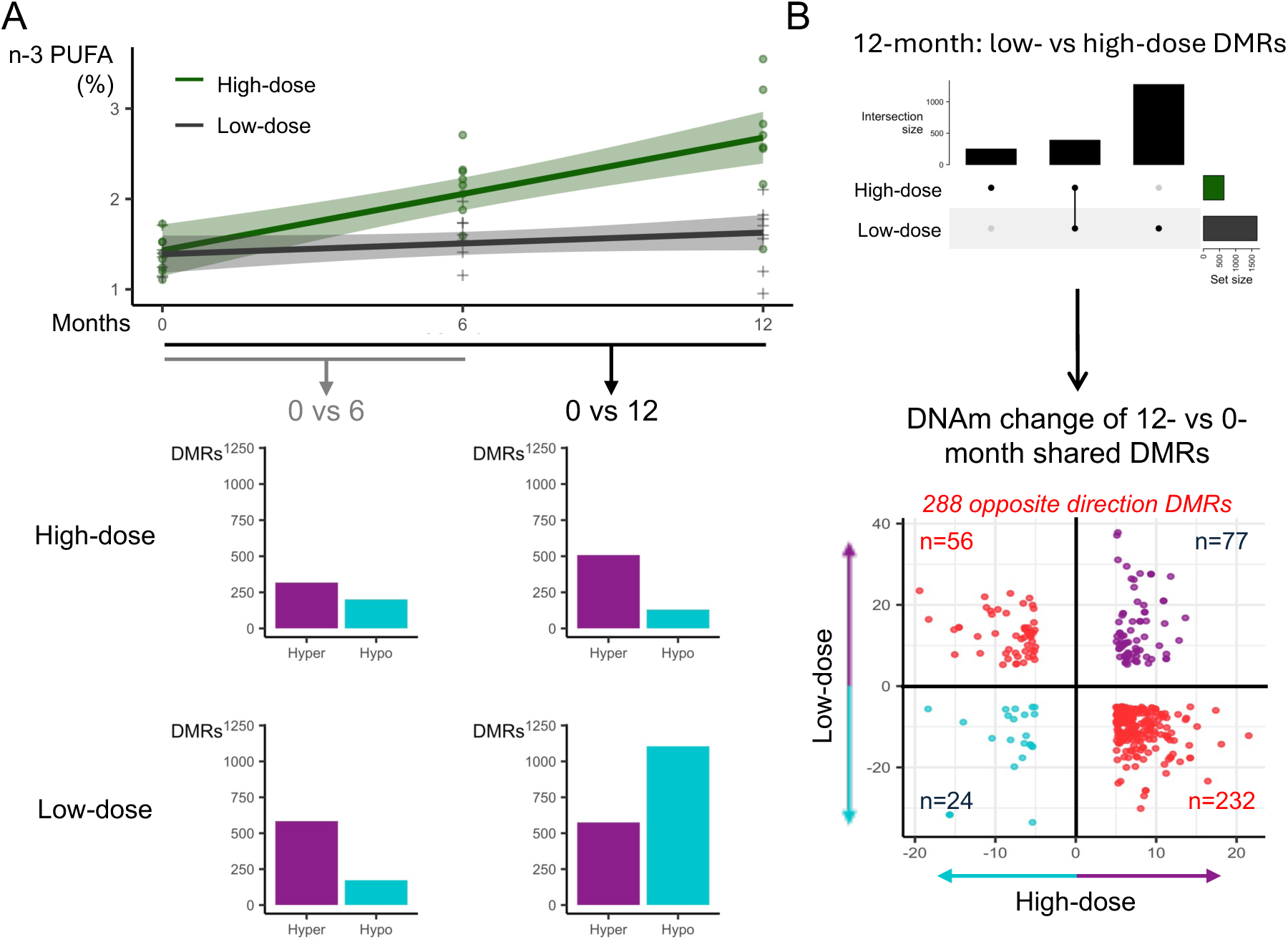
Differential methylated regions (DMRs) of high- and low-dose over time. **A)** Plots depict DMRs (bottom) and n-3 PUFA changes in breast adipose (top) by dose arm (n=7 high-dose, n=10 low-dose participants) over time (0, 6, 12 months). The change in total n-3 PUFAs is shown for each dosing arm using linear fit lines and 95% confidence intervals (top). Gene promoter DMRs were classified as either hypermethylated (DNAm increased after treatment) or hypomethylated (DNAm decreased after treatment) for each dosing arm (bottom). **B)** The upset plot summarizes the intersection between the baseline and 12-month DMRs (top). 389 DMRs were shared between high- and low-dose. The scatter plot (bottom) depicts the direction of DNAm change in each dosing arm for the 389 shared DMRs, the majority (74%) changing inversely between arms (bottom; red points in quadrants 2 and 4).

### DNAm changes show hypomethylation in immune pathways

By pathway enrichment analysis of 0- to 12-month DMRs, we explored the specific biologic processes affected by DNAm changes following high-dose n-3 PUFA treatment (**Fig. 3A**). The high-dose arm DMR analysis identified seven significantly enriched pathways involving hypomethylated DMRs associated with immune-related processes (**Table S1**). Interestingly, the hypermethylated DMRs, which were more numerous than the hypomethylated DMRs, were not significantly enriched in any pathway. Additionally, we attempted to identify enriched processes using different comparison strategies, pathway databases, and genomic regions (**Fig. S4**). These additional strategies produced similar results with low numbers of hypomethylated DMRs in immune-related processes and no enrichment for hypermethylated DMRs (**Fig. S4; Table S2**).

**Figure 3.**
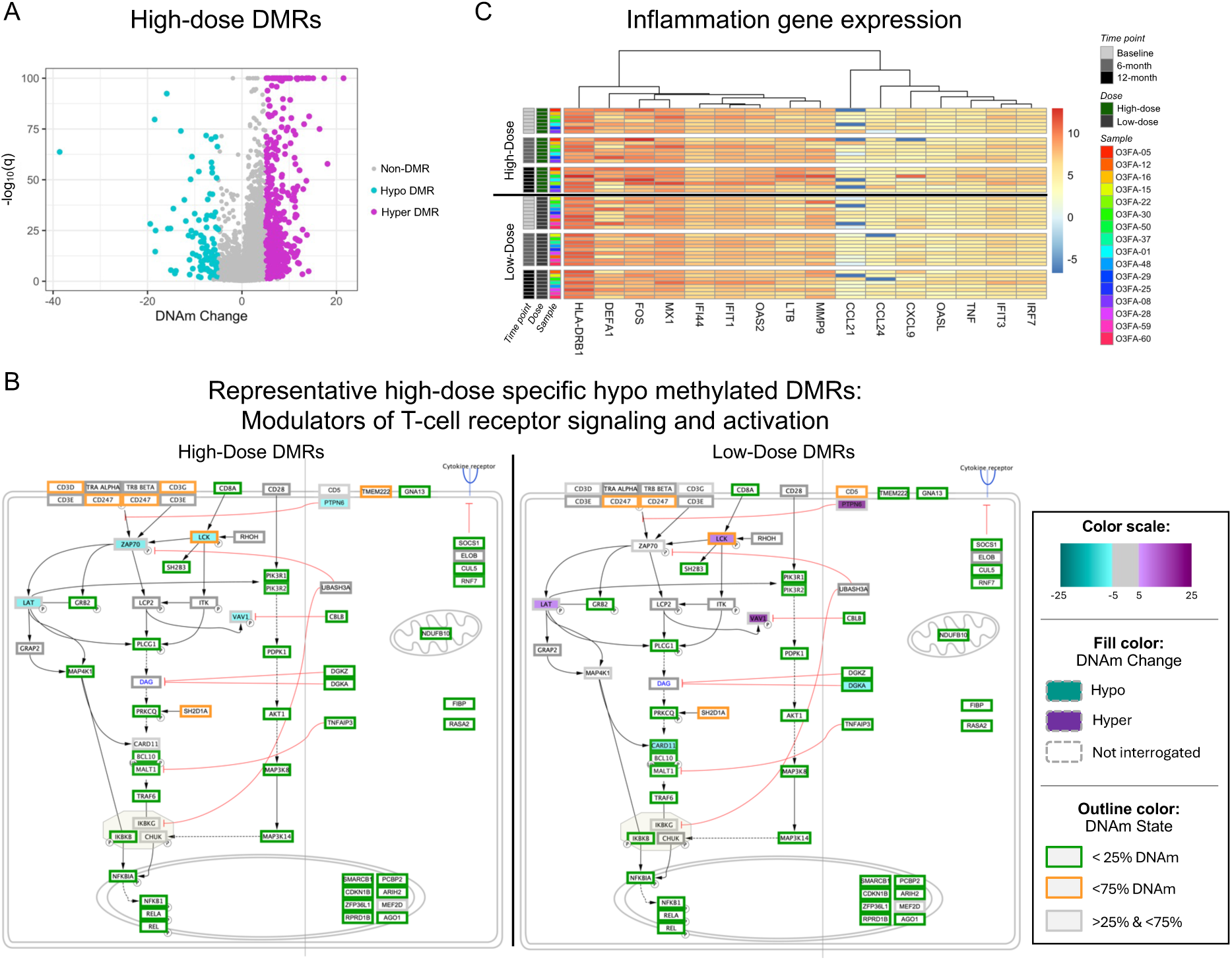
Pathway analysis of DMRs and inflammation related expression changes. **A)** A volcano plot of the high-dose promoter DMRs shows the magnitude of DNAm changes. **B)** DNAm change of the DMRs for both the high-dose (left) and low-dose (right) in the T-cell receptor signaling pathway, the most significantly enriched pathway from the hypomethylated high-dose DMRs. The low-dose DMRs showed no enrichment for any pathways. **C)** Gene expression for all genes differentially expressed in either the high- or the low-dose group. Gene expression assayed using a NanoString nCounter panel that assessed 249 inflammation-related genes.

Conversely in the low-dose arm, despite the larger number of DMRs, neither the hypo- nor the hyper-methylated DMRs were significantly enriched in any pathways. When we looked at the high-dose arm DMRs that were significantly enriched in immune-related pathways, all five of these genes had an opposite DNAm change (hypermethylated) in the low-dose arm, as exemplified by the T-cell receptor signaling and activation pathway (**Fig. 3B**). Overall, pathway enrichment analysis identified very few high-dose DMRs (5 of 638; **Table S1**) and no low-dose DMRs that were enriched in biological pathways.

### n-3 PUFA treatment is associated with heterogeneous increases in inflammation-related gene expression

To explore inflammation-related gene expression changes in breast adipose tissue induced by high- and low-dose n-3 PUFA supplementation, we assessed breast adipose tissue from the same time point (83% replicates with the RRBS samples) for expression of inflammation-related genes (249 gene NanoString nCounter Gx Human Inflammation). For each dosing arm, differentially expressed gene (DEG) analysis of the 0- to 6-month and 0- to 12-month time points identified DEGs in only the 0- to 12-month comparisons (**Fig. 3C**). In the high-dose arm, sixteen genes were identified as DEGs with increased expression after n-3 PUFA treatment in all but one gene (**Fig. 3C**; **Table S3**). However, all DEGs had variable levels of expression between individuals and expression changes that were small in magnitude (average log2 foldchange =1.1), Five of the high-dose DEGs were also DEGs in the low-dose arm, and only one DEG was unique to the low-dose arm (**Table S3**).

### Breast adipose DNAm promoter heterogeneity is reduced following high-dose n-3 PUFAs

Given the divergent DNAm responses in the high- versus low-dose arms, in terms of global DNAm (CpG-based distribution) variance (**Fig. 1B; S2B-C**) and DMR direction of change (**Fig. 2B; 3B**), we suspected that n-3 PUFA treatment might alter epigenetic (DNAm) fidelity in addition to the steady-state DNAm levels, utilizing both information-theoretic (i.e., entropy) and variance-based DNAm assessments.^30,31^ Entropy-based methods quantify the distribution of DNAm at a genomic locus to capture stochastic failures of DNAm maintenance which has been linked to both aging and breast cancer.^32,33^ Heterogeneous distribution of DNAm produces high entropy values whereas a homogeneous or uniform distribution of DNAm produces low entropy values. For the entropy-based approaches, we found low entropy in both arms (**Fig S5A**); the low entropy may relate to the RRBS assay which interrogates CG-dense, promoter associated regions more likely to be strongly regulated and therefore with more uniform DNA.^33^

Variance-based approaches have been used previously to detect stochastic epigenetic field defects in normal breast tissue adjacent to cancer. These epigenetic changes were identified as critical events in early epigenetic transformation and may prove well-suited to detecting changes from n-3 PUFA treatment.^19,34^ To identify changes in DNAm variance, we tested all CpGs for changes in DNAm variability by comparing baseline versus 12-month treatment samples in each arm.^19^ After identifying differentially variable (DV) CpGs, we found that breast adipose tissue of the high-dose arm was enriched for decreased DNAm variability while the low-dose arm tissue was enriched for increased DNAm variability (p<0.001; **Fig. 4A; Fig. S5B**). For DV CpGs identified by comparing dosing arms (high- vs low-dose), there was no change in variability at baseline but decreased DNAm variability in the high-dose arm at 12 months (**Fig. S5C**). The majority of DV CpGs in both arms were identified in low DNAm (<25% DNAm) CpGs (**Fig. 4B; Fig. S5B**). Additionally, the DV CpGs were enriched in promoters and significantly depleted in DMRs in both dosing arms (**Fig. 4C**; **S5D**). The exclusion of DV CpGs in DMRs suggested that changes in DNAm variability are independent from the changes in steady-state DNAm (i.e., DMRs). Notably, the DV analysis of PBMC DNAm using 0 and 6 months of high-dose (5 g EPA+DHA/d) n-3 PUFA supplementation^35^ demonstrated a similar trend for decreased DNAm variability after n-3 PUFAs (**Fig. S5E**). Together, the observed changes in DNAm variability may indicate that high-dose n-3 PUFA supplementation is associated with stabilized or increased DNAm fidelity whereas the low-dose arm is associated with a loss of DNAm fidelity.

**Figure 4.**
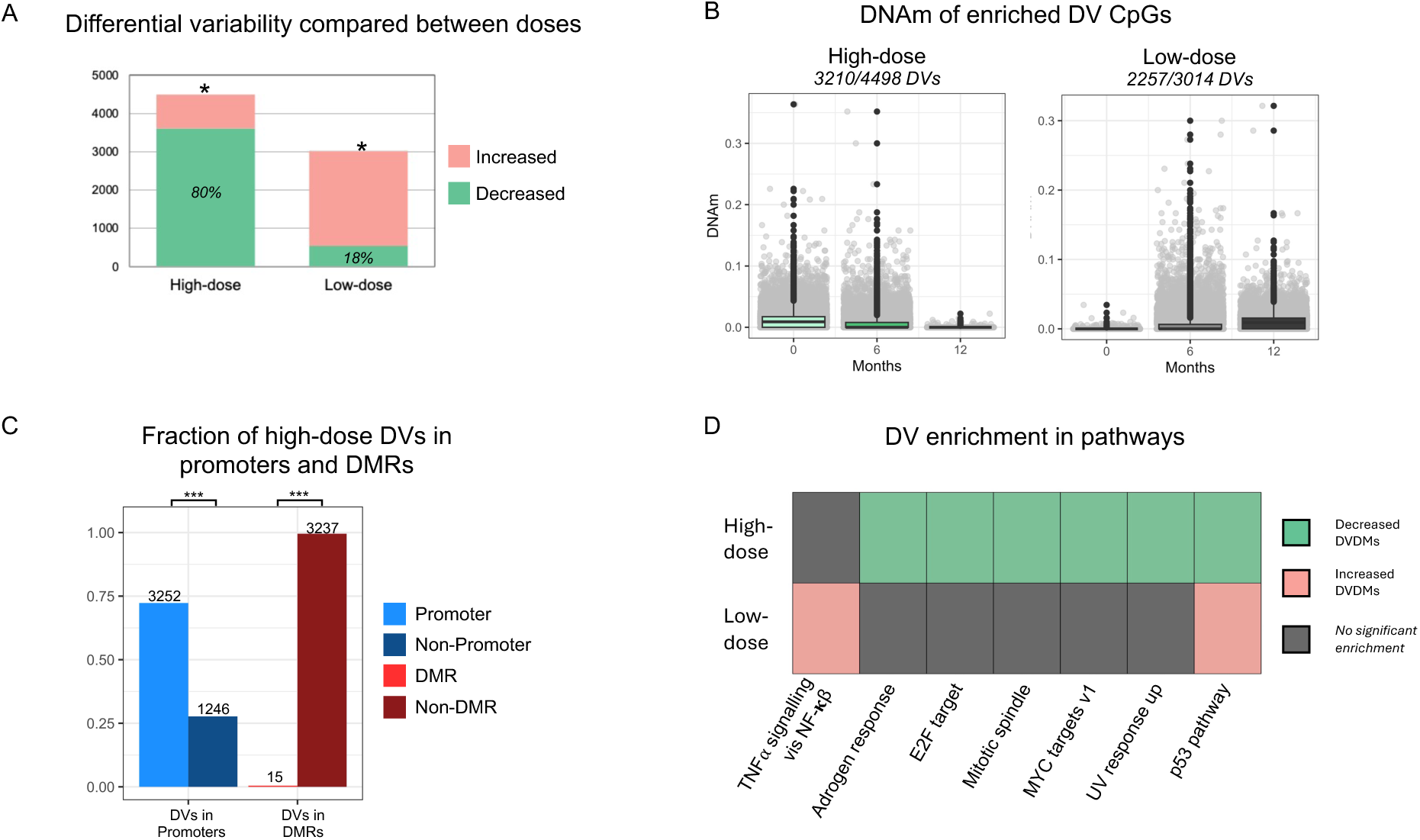
n-3 PUFA effects on DNAm variability. **A)** The number of identified DVDMs for each treatment arm. Hypergeometric tests determined enrichment of decreased variability DVDMs in the high-dose and increased variability DVDMs in the low-dose arm (*pval<0.001). **B)** DVDMs are depicted by DNAm level (high vs low) and variability change (increased vs decreased), with DNAm plotted to visualize the DNAm variance change for combination that had the highest number of DVDMs in each dosing arm (see **Fig. S5D** for DNAm changes of all four combinations in each dosing arm). **C)** DVDMs intersected with both the promoter regions and DMR promoter regions show significant enrichment of DVDMs in the promoters but significant depletion in the DMRs (***hypergeometric pval <0.0001). **D)** DV enrichment was performed using multiple pathway and gene set databases (**Table S4; Fig. S6**), with the trends and types of biological processes significantly enriched by the high- and low-dose DVs summarized using the mSigDB Hallmark gene set enrichment.

To assess the potential impact of the DVs on biological processes, we performed a pathway enrichment analysis on all baseline vs 12-month DVs. Using multiple pathway and gene set databases, we assessed enrichment of the increased vs decreased DVs separately for both the high- and low-dose arms (**Table S4**). Consistent with the directional trends of the DVs identified in each arm, we found that the significantly enriched pathways were primarily from decreased DVs in the high-dose arm and increased DVs in the low-dose arm. Additionally, most significantly enriched pathways were unique to either the high-dose or the low-dose arms. In particular, the mSigDB Hallmark gene set^36^ showed largely distinct pathway enrichment between the high- and low-dose arm and only one shared pathway (**Fig. 4D**). Similar to the enrichment observed in the Wikipathway database used for DMR enrichment (**Fig. S6**), Hallmark gene set pathway analysis shows DV enrichment predominantly in processes related to oncogenic signaling and genome stability.

## DISCUSSION

With this sub-study of breast adipose DNAm in ERPR(−) breast cancer survivors on high-dose versus low-dose n-3 PUFA supplementation for 12 months, we demonstrated dose-specific DNAm effects. While both dosing arms exhibited treatment-specific DNAm changes, only the high-dose arm showed DNAm enrichment for inflammation-related pathways (**Fig 3B; S4X**). We also determined that DNAm variability decreased with high-dose but not low-dose n-3 PUFA treatment, which suggests a dose-dependent treatment-induced change in epigenetic fidelity in breast adipose tissue in this study cohort of ERPR(−) breast cancer survivors.

Notably, the relationship between n-3 PUFA dose and breast adipose DNAm was complex, differing from the dose-dependent increases in the n-3 fatty acids EPA and DHA and derived metabolites (**Fig. 5**). Significant differences in breast adipose DNAm patterns between the high-and low-doses included: 1) many of the DMRs shared by both study arms change in the opposite direction (**Fig. 2B**); 2) the low-dose group had an overall greater number of DMRs (**Fig. 2A**); and 3) DNAm variability decreased in the high-dose group yet increased in the low-dose group (**Fig. 5A**). These changes in adipose DNAm are directionally discordant and dose-dependent in a non-linear fashion, with quantitatively distinct epigenetic responses elicited by high- and low-dose n-3 PUFA supplementation. Together, these data raise the possibility of bifurcation-like behavior in n-3 PUFA dose and DNAm effects in breast adipose tissue: in the breast adipose n-3 PUFA crosses a critical dose threshold that shifts the system between two alternative DNAm states and produces directionally opposite DNAm changes and DNAm variability.

**Figure 5.**
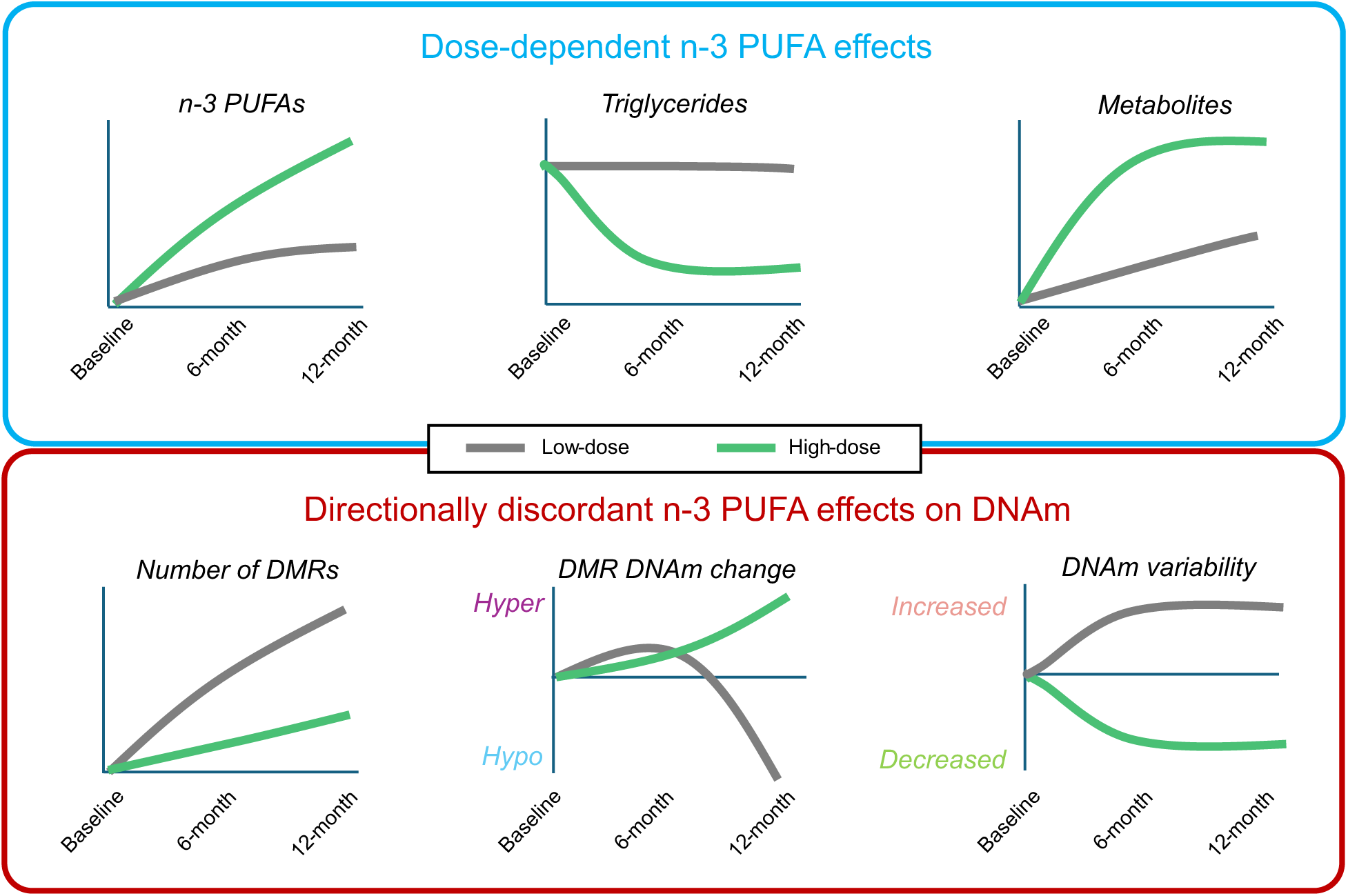
Graphical summary of high- vs low-dose n3-PUFA effects. Graphs depict the n-3 PUFA dose-dependent (higher dose ∼ larger effects, top panel) effects for breast adipose total n-3 PUFAs and oxylipin metabolites and serum triglycerides and directionally discordant (bottom panel) effects in breast adipose tissue DNAm for DMR DNAm change and DNAm variability (n=7 high-dose, n=10 low-dose participants). The total number of DMRs (i.e., steady state DNAm changes due to n3-PUFAs) were higher in the low-dose, and both the direction of DNAm change in the DMRs and the DNAm variability changed in opposite directions between the high-dose and low-dose n3-PUFA arms.

The change in DNAm variability observed in our study has important implications for n-3 PUFA supplementation and breast cancer prevention. Increasing evidence links DNAm variability with cancer development and progression that may reflect environmental or treatment effects.^37–40^ Prior studies have also demonstrated DNAm variability in adjacent normal tissue as a field effect that precedes breast cancer development.^19,34^ Teschendorff et al.^19^ showed that DVs identified in normal tissue adjacent to breast tumors were enriched for cancer related pathways. Here, we demonstrated that DVs in breast adipose tissue were enriched in many different biological processes including cancer-related pathways. Our low-dose arm had increased DNAm variability similar to the results of Teschendorff et al. as field effects in breast cancer; however, the high-dose treatment reduced DNAm variability. As survivors of ERPR(−) breast cancer, study participants might be at risk of a cancer-prone mammary microenvironment with accumulating field defects in the form of dysregulated DNAm even if disease-free for the duration of the trial. In this scenario, the low-dose treatment may have been insufficient to offset DNAm dysregulation, thereby yielding increased DNAm variability over the 12-month intervention, whereas high-dose n-3 PUFAs may have restored DNAm fidelity by suppressing dysregulation which resulted in decreased DNAm variability. With increasing evidence linking cancer risk and disease progression to highly variable DNAm, the inhibitory effects of high-dose n-3 PUFAs on DNAm variability in the breast adipose microenvironment may represent a novel mechanism of cancer prevention. Based on our study findings, we hypothesize that high-dose n-3 PUFA supplementation has two protective effects on DNAm in the breast microenvironment of women at high-risk: 1) a small number of enriched biological processes which we observed as DNAm changes in gene promoters, and 2) inhibition of dysregulation of DNAm that restores DNAm fidelity in the breast environment.

Previous n-3 PUFA studies, in the context of breast cancer and otherwise, have reported both anti-inflammatory effects and DNAm changes after n-3 PUFA treatment.^41^ In our sub-study of DNAm in PBMCs from women at high risk of breast cancer comparing high- vs low-dose n-3 PUFA, we observed DNA hypermethylation in inflammation-related pathways (Toll-like receptor signaling) at 6 months.^35^ In the current study of breast adipose DNAm at 0, 6, and 12 months of ∼1 g EPA+DHA/d versus ∼5 g EPA+DHA/d, we observed hypomethylation in a different set of inflammation-related pathways such (i.e., T-cell receptor and Interferon-*γ* signaling). Prior research has also demonstrated distinct DNAm patterns by tissue type as with blood and breast tissue.^42^ By NanoString technology, we observed small and heterogeneous changes in expression of inflammation-related genes in breast adipose tissue samples (**Fig. 3C**). However, in matched breast adipose samples, we previously reported significantly increased EPA- and DHA-derived oxylipin fatty acid metabolites which may elicit pro-resolving, anti-inflammatory effects and may more accurately represent the anti-inflammatory effects of n-3 PUFAs.^28,43^

Limitations include the sub-study sample size of 17 of 51 participants who completed the 12-month intervention due to constraints of funding and resources. Although designed to sample breast adipose tissue for a read-out of microenvironment inflammation or metabolic dysregulation before and after dietary n-3 PUFAs, tissue collection by fine needle aspiration (FNA) will reflect the mixed cell population of adipose tissue, including immune, endothelial, and stromal-vascular cells in addition to adipocytes and pre-adipocytes as the major components. Nonetheless, our analyses of breast adipose FNA samples demonstrate highly significant differences in microenvironment DNAm before and after high-dose n-3 PUFAs. Future research using single cell technology may further elucidate cell-type-specific signaling mechanisms.^44,45^ Analysis of DNAm entropy was limited by RRBS methodology, which relies on highly regulated CpG dense regions, and further research using array-based technology will enable investigation of DNAm entropy in the uninvolved, at-risk breast tissue microenvironment. Moreover, we found that only a small fraction of the DMRs (5 of 408 total) were enriched in biological pathways such that the biological effect of the majority of DNAm changes remain undefined. Future studies that pair unsupervised expression experiments with DNAm would both identify the general effects of n-3 PUFA on gene expression and better resolve the biological role of the large number of DNAm changes that were not attributed to biological pathways.

Taken together, our findings show that different doses of n-3 PUFAs result in quantitatively distinct and multi-faceted effects on DNAm. The complex, directionally discordant effects of high-relative to low-dose n-3 PUFAs on both steady-state DNAm and DNAm fidelity reveals important considerations for designing future n-3 PUFA interventions. Importantly, the finding that high-dose n-3 PUFA affects DNAm fidelity in the breast adipose suggests a new potential mechanism for n-3 PUFA-mediated prevention of breast cancer development.

## METHODS

### Study design and participants

This study was a substudy of a randomized, dose-comparison clinical trial of long-chain n-3 PUFA supplementation (high-dose vs low-dose) in women at elevated risk for hormone-receptor-negative breast cancer (*Clinicaltrials.gov:* NCT02295059). The substudy included a subset of the participants for whom breast adipose fine-needle aspirates (FNA) were available at baseline (0 mo), 6 mo and 12 mo. Baseline characteristics, eligibility criteria, randomization procedures, and sample handling followed the parent clinical trial protocol.^28^

### Sample collection and storage

Breast adipose tissue was obtained by ultrasound-guided fine-needle aspiration (FNA) at each study visit and immediately processed according to the parent trial protocol. Samples were stored at −80 °C until DNA extraction. To adjust for sampling heterogeneity, lipid-mediator concentrations (including 13-HODE) were measured on matched aliquots and were included as covariates in downstream models where indicated.

### DNA extraction and RRBS library preparation

Genomic DNA was extracted from FNA material using the Gentra Puregene DNA extraction kit (Qiagen). Samples were required to yield at least 50 ng genomic DNA for library preparation. RRBS libraries were generated using a modified reduced-representation bisulfite sequencing protocol adapted from the parent trial as follows: MspI was used to digest genomic DNA at 37 °C overnight. End repair and 3′ A-tailing were performed in the same tube using Klenow Fragment (3′→5′ exo-minus), followed by ligation to methylated universal adapters (IDT xGen™ Stubby Adapter, Cat. 10005924). Ligation products were purified with 2.0× Ampure XP beads (Beckman Coulter, Cat. A63881) and subjected to bisulfite conversion using the EZ DNA Methylation kit (Zymo Research, Cat. D5001). Final libraries were amplified with Pfu Turbo Cx Hotstart DNA Polymerase (Agilent, Cat. 600414) using IDT xGen™ UDI primers (Cat. 10005922) and purified with 1.2× Ampure XP beads. Library quality control was performed on an Agilent TapeStation D5000 and libraries were quantified by qPCR. Libraries were sequenced on an Illumina NovaSeq 6000 instrument (S4 reagent kit v1.5, 300 cycles). Base calling was performed by Real-Time Analysis (RTA) v3.4.4 and BCL files were converted to FASTQ using bcl2fastq (RRID:SCR_015058). A minimum of 40 million reads was sequenced for each library (mean total reads: 64 million).

### Read processing and alignment

Raw reads were processed through a reproducible nf-core-methylSeq v1.6.1 pipeline. Adapter trimming and low-quality base filtering were performed prior to alignment. Trimmed reads were aligned to the human reference genome (GRCh38/hg38) using Bismark (version 0.23.0) and the HISAT2 (RRID:SCR_015530) aligner (version 2.2.1) and methylation calls were extracted using the Bismark methylation extractor (methXtract). CpG coverage files and per-CpG DNAm β-values were generated for downstream analyses. All pipeline runs were containerized and executed in a cluster environment to ensure reproducibility. Per-sample QC metrics included total read count, mapping rate, duplication rate, bisulfite conversion efficiency, and per-CpG coverage distribution. All samples passed QC, and the incomplete conversion rates were low (mean=.99%, max=1.55%). CpG sites were retained for analysis only if covered by at least 10 reads in the sample set used for a given comparison. Global CpG methylation distributions and principal component analysis (PCA) were inspected to detect outliers and batch effects.

### DNAm summarization and promoter definitions

Promoter regions were defined as ±1 kb around annotated transcription start sites (TSS) from the RefSeq hg38 annotation. CpG islands genomic coordinates were defined by the UCSC genome browser hg36 track. The RefSeq gene annotations were also used to define the 1^st^ exon and gene body genomic features (TSS to transcriptional end site). An additional long-range distal promoter was defined as 10kb up- and 1kb down-stream of the RefSeq TSS, and the Promoter associated CpG Islands were defined as any CpG island that had any intersection with a distal promoter region. For each genomic feature, DNAm was summarized as the mean β-value across all CpGs that met the 10 read coverage criteria within each region.

### Differentially methylated region (DMR) analysis

DMR detection was performed using methylKit to identify genomic regions with significant changes in mean DNAm between time points and between arms. Only CpGs that had >10 reads were included in a region and a DMR was defined as a region that had both a multiple testing corrected false discovery rate (FDR) p < 0.05 and at least a 5% average DNAm difference which was set because it was >2x the maximum incomplete conversion rate detected. Using the average DNAm within each group, DMRs were labeled with respect to the baseline DNAm as either hypermethylated (increased average DNAm after treatment) or hypomethylated (decreased average DNAm after treatment).

### Differential variability analysis (iEVORA)

CpG-level tests for changes in inter-sample variability were performed using the iEVORA algorithm to identify differentially variable CpGs (DVs) between baseline and post-treatment samples within each arm using the iEVORA R package with the default parameters for calling a CpG significantly variable.^19^ DVs were associated with promoter and DMRs by intersecting their genomic coordinates using the previously described promoter/region definitions. Enrichment analyses were performed to test whether DVs were over- or under-represented in promoters or DMRs using the hypergeometric test.

### Entropy and epiallele diversity analyses

Epiallelic entropy was computed following the approach of Jenkinson et al. (2017) to quantify the diversity of methylation patterns at contiguous CpG sites (DNAm entropy).^33^ Entropy calculations were restricted to loci with sufficient contiguous, read-level coverage to permit reliable epiallele reconstruction. Entropy was calculated both horizontally (within sample diversity) or vertically (intersample diversity) as both Shannon and normalized Shannon entropy *H*(*x*) = − ∑ *p*(*x*)*log*_!_(*p*(*x*)) where *p*(*x*) is the probability of a CpG being methylated as *x* ranged over either the horizontal (within a read) or vertical (between reads) DNAm values of each CpG site.^33^ Since entropy was uniformly low in both arms and at all time points for this RRBS dataset, we interpreted that as a consequence of the CG-dense, promoter-associated regions preferentially interrogated by the RRBS assay and therefore deprioritized entropy as the primary heterogeneity readout for the main set of analyses.

### NanoString targeted gene expression assay

A targeted inflammation gene panel (Gx Human Inflammation; n = 249 genes) was assayed on matched breast adipose samples using the NanoString nCounter platform. RNA was extracted, hybridized and counted according to the manufacturer’s instructions. Raw counts were normalized following NanoString recommended procedures (positive control normalization, code-count normalization, and housekeeping gene scaling) and were analyzed for differential expression using the NanoStringNorm (version 1.2.1) and DESeq2 (version 1.44.0) R packages.

### Pathway and gene set enrichment analyses

Genes annotated to promoter DMRs, DV CpGs or DVDMs were tested for enrichment usin the enrichR (version 3.2) R package against curated gene sets including Wikipathway (WikiPathways_2024_Human), MSigDB Hallmark (MSigDB_Hallmark_2020), MSigDB oncogenic signatures (MSigDB_Oncogenic_Signatures), KEGG (KEGG_2021_Human), and Reactome (Reactome_Pathways_2024).^46^ For all comparisons, statistical significance was defined as an adjusted p-value < 0.05.

### External validation and comparative cohorts

DV directionality was assessed in an independent peripheral blood methylation dataset.^35^ DVs were called using the iEVORA-based same procedure and criteria for significance described previous. The effects of n-3 PUFAs on PBMCs were compared using baseline versus 6-month end of study samples obtained from participants in the 5 g/d EPA+DHA arm of a randomized dose-finding trial of n-3 PUFAs in women at high risk of breast cancer.^47^

### Statistical rigor and reproducibility

Statistical analyses were designed and executed to maximize rigor, limit bias, and ensure reproducibility. All analyses were run using containerized pipelines and scripted R workflows so that every step (read processing, methylation calling, DMR/DV and downstream enrichment tests) can be rerun identically. The primary processing pipeline was executed in a containerized nf-core/RRBS environment and downstream analyses were performed in R (versions and package versions are provided). We applied a minimum per-CpG coverage threshold of 10 reads for inclusion in analyses and a minimum threshold for defining DNAm differences based on the maximum observed incomplete conversion rate to control for false postives. All hypothesis tests were two-sided, and to control the false discovery rate from genome-scale testing, multiple test correction was reported at FDR < 0.05 (unless otherwise noted); exact p-values, effect sizes (absolute methylation differences) and where appropriate 95% confidence intervals are reported.

### Code and data availability

The code and custom analysis scripts used to produce all analysis and figures are available at (*github link forthcoming*). Raw sequencing data (FASTQ) and processed methylation matrices are available via Gene expression omnibus (*GEO accession pending*).

## Data Availability

All data produced will be available online (GEO accession pending)

## Acknowledgments

We gratefully acknowledge the time, effort, and commitment of the study participants. We thank all the clinic staff, clinicians, and clinical trials team involved in this study at City of Hope National Medical Center/COHCCC and Stefanie Spielman Comprehensive Breast Center/OSUCCC. We acknowledge Dr Joanne L Lester and Ms Maria V Johnson for invaluable assistance with the conduct of the clinical trial and data management.

## Funding

This clinical trial was funded by R01 CA164019 (LDY). We are grateful for the COHCCC Core Facilities supported by NIH P30CA033572, including the Integrative Genomics Core. Additional support was provided by USDA Project 2032-51530-025-00D (JWN). The USDA is an equal opportunity employer and provider. The USDA had no influence in the study design or interpretation of the experimental results. The reported findings and their interpretation are the expressed opinions of the authors and do not represent an official position of the USDA.

## Supplementary Figures

**Figure S1.**
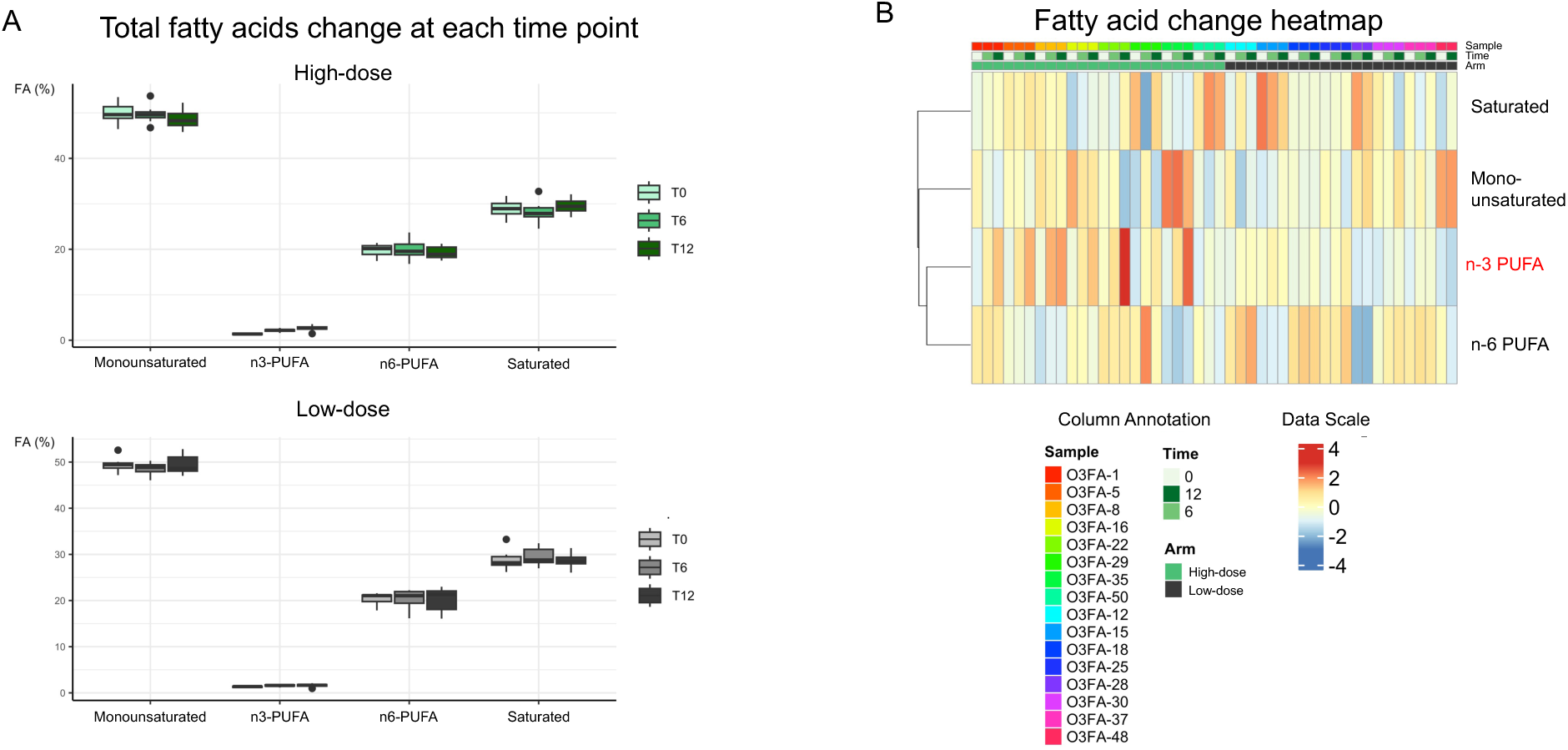
Fatty acid data for the DNAm substudy breast adipose tissue samples. **A)** Summarized fatty acid changes over time in both the high- and low-dose arms for the samples included in this sub-study. **B)** Fatty acid values for each sample were shown in a heatmap to visualize both changes and heterogeneity.

**Figure S2.**
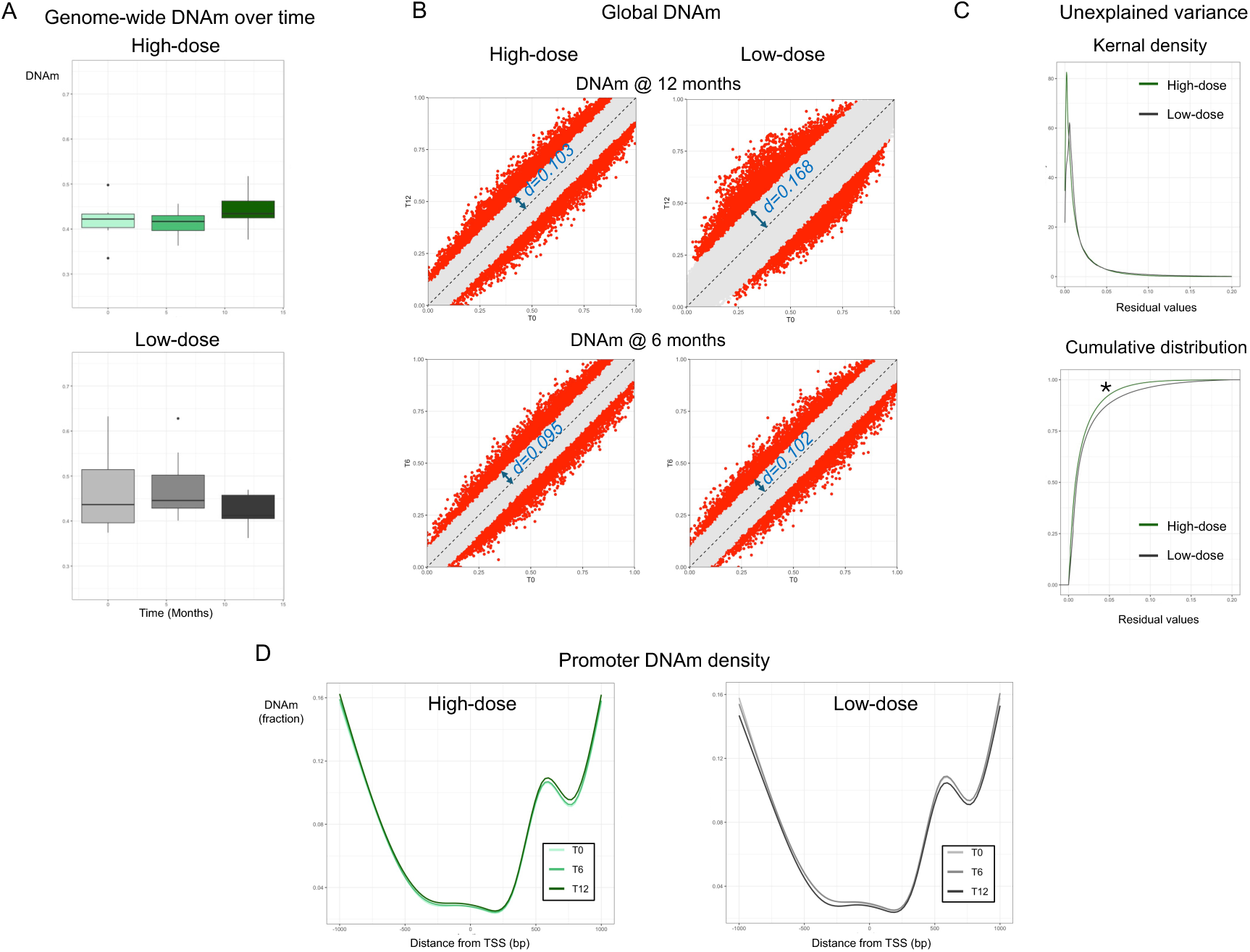
**A)** Global DNAm changes over time in both treatment arms (n=7 high-dose, n=10 lowdose particpants with 0-, 6-, 12-month samples). **B)** Comparison of global DNAm variance between high- and low-dose groups by measuring the width of the 99th percentile bands. CpGs outside the 99th percentile are shown in red. **C)** Comparison of the unexplained DNAm variance between treatment arms by plotting the distribution of the residuals of the fit for baseline vs 12-month global DNAm. The unexplained variance is depicted both as the density of residuals (left) and as a cumulative distribution function (right; pval< 0.001). **D)** Summary of DNAm 1kb up- and down-stream of the transcription start site (TSS; bp=0) for each treatment arm and at each time point to assess the genomic distribution of DNAm in all gene promoters and determine treatment effects on the distribution of DNAm around TSS.

**Figure S3.**
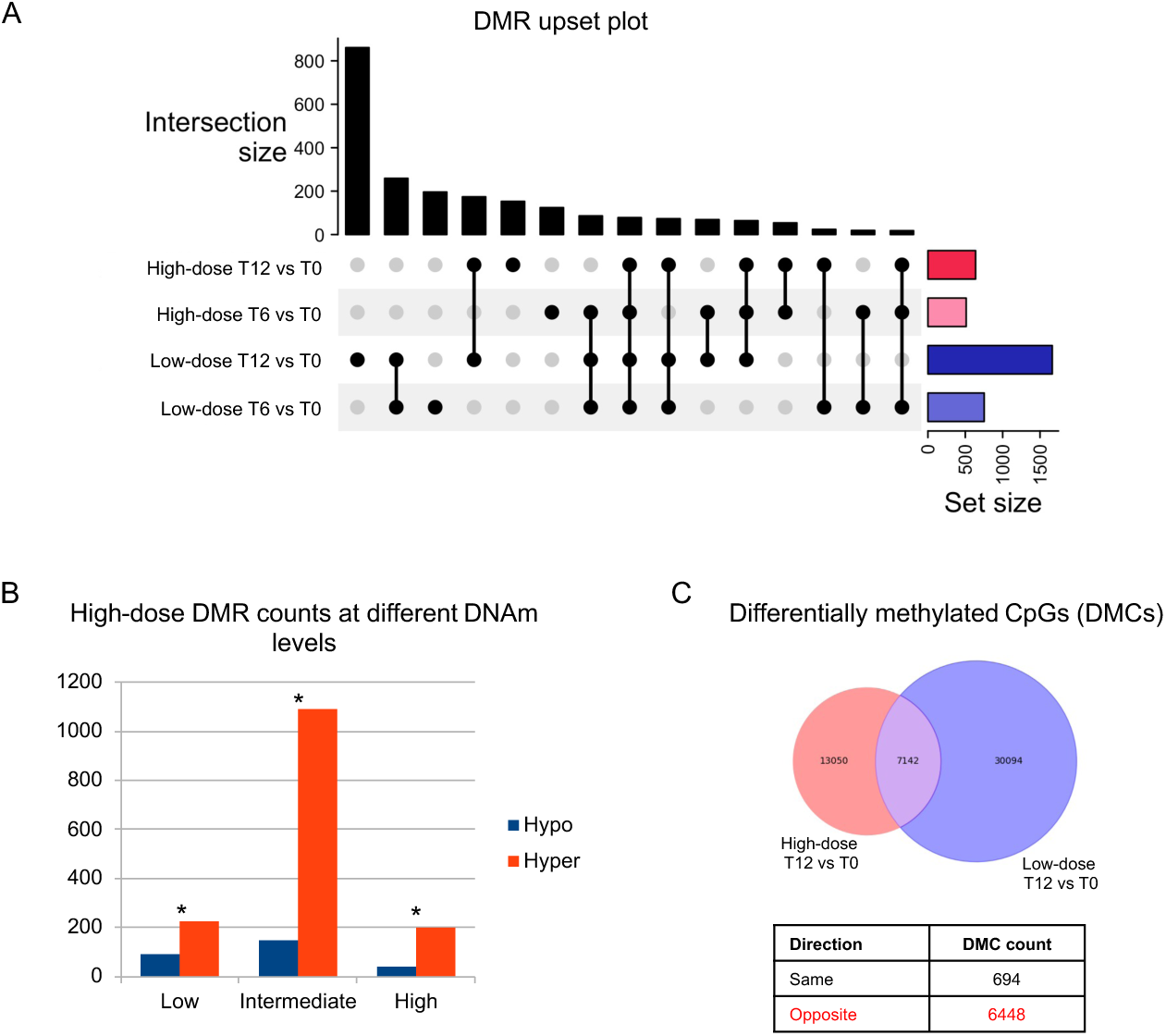
**A)** Upset plot of the total number of DMRs and all intersections of the high- and low-dose DMRs with comparisons between both 6- and 12-months vs baseline. **B)** The number of DMRs from the high-dose 12-month vs baseline comparison shown by direction of DNAm change at three levels of DNAm. DNAm was stratified using 12-month DNAm into low (<25%), intermediate (>25% and <75%), and high (>75%) DNAm for hypomethylation (Hypo) and hypermethylation (Hyper). Hypergeometric testing showed enrichment for hypermethylation at all levels of DNAm (p < 0.05). **C)** Differentially methylated CpG analysis confirmed the DMR results that most of the methylation changes were shared between the high- and low-dose arms change in the opposite direction.

**Figure S4.**
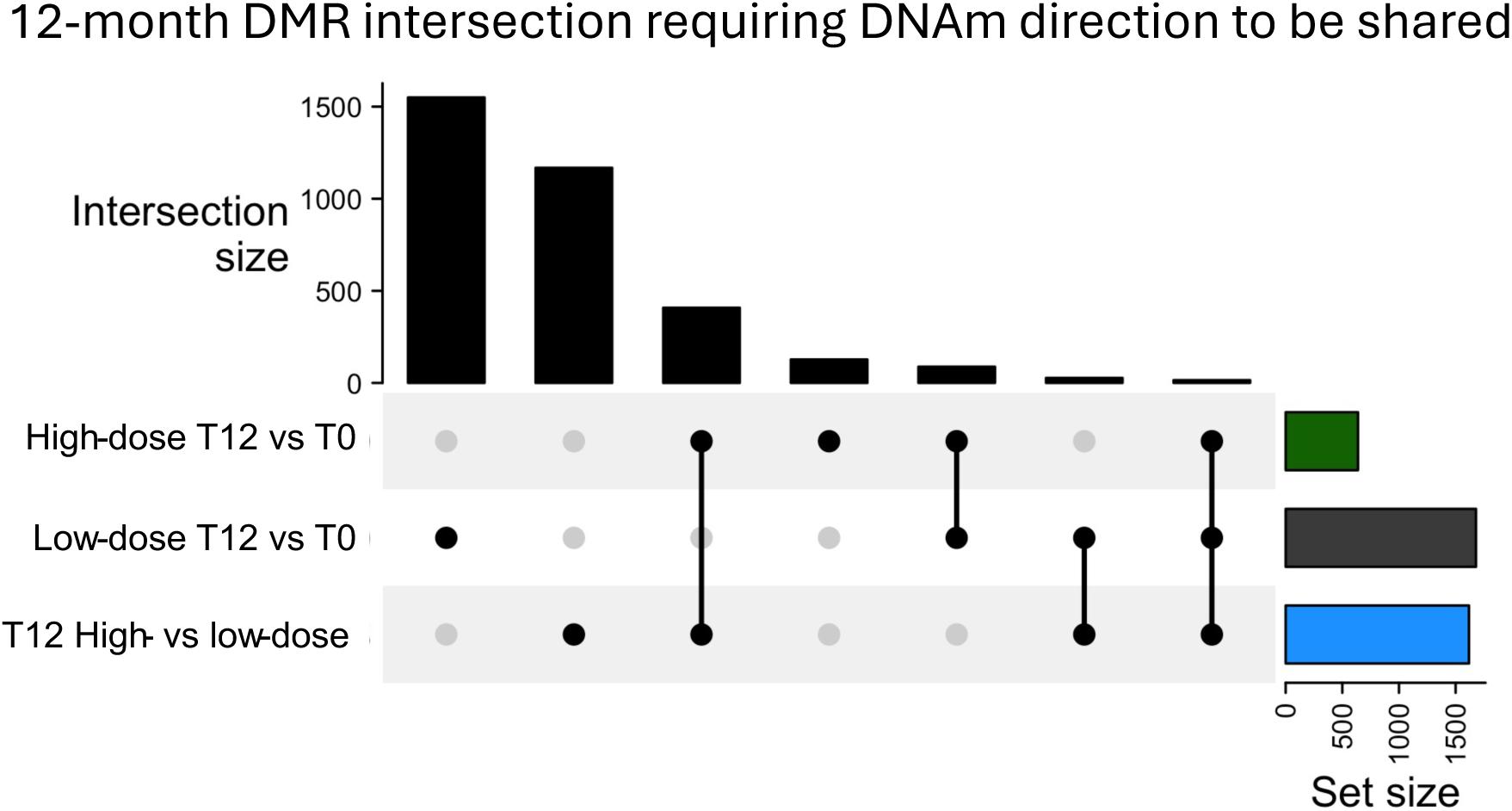
Upset plot of the intersection of the 12-month DMRs for both arms and the DMRs that were identified by directly comparing the 12-month high-dose to the 12-month low-dose samples.

**Figure S5.**
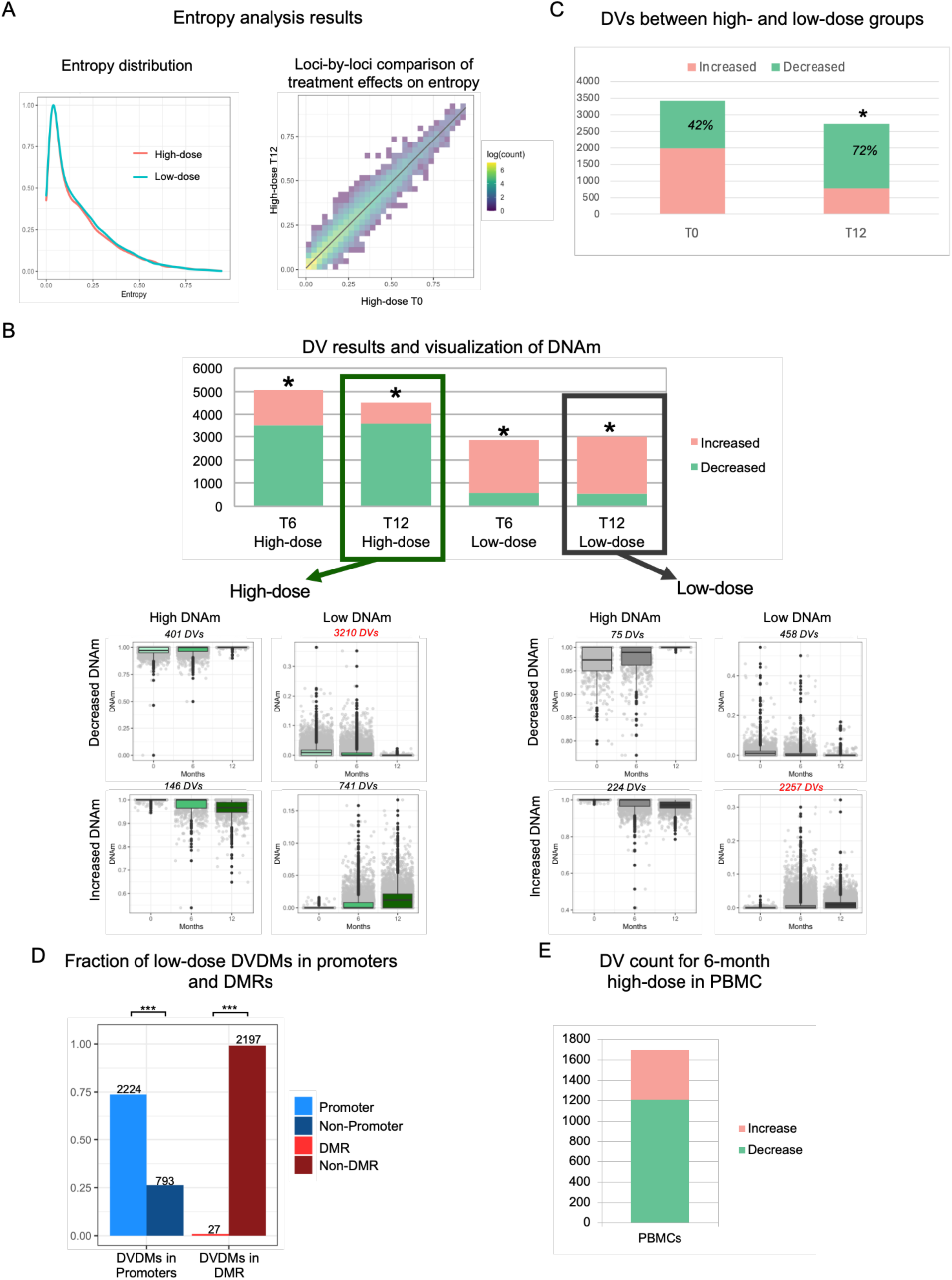
**A)** Entropy analysis on RRBS data showed low entropy in both arms (left) and no differences after high-dose treatment (right). **B)** DV CpGs were identified by directly comparing baseline to both the 6-month and 12-month time points in each dosing group (top). By hypergeometric test the high-dose group was significantly enriched for decreased DVs while the low-dose group was enriched for increased DVs (* p<0.01). DVs are depicted by plotting DNAm of all DVs based on the DNAm level (low vs high) and the direction of change in variability (increased vs decreased; bottom). The total number of DVs in each plot are indicated. **C)** Direct comparison of DV CpGs at baseline and 12-months for high- vs low-dose groups (* hypergeometric test p<0.01). **D)** Similar to the high-dose DV CpGs, the low-dose DV CpGs were significantly enriched in the promoter region and significantly depleted in the promoters identified as DMRs (*** hypergeometric p < 0.0001) **E)** The trend for decreased variability DVs observed in the high-dose n-3 PUFA were validated an independent cohort of PBMCs collected from women treated with high-dose n-3 PUFAs (5.04 g EPA+DHA/d) for 6-months.

**Figure S6.**
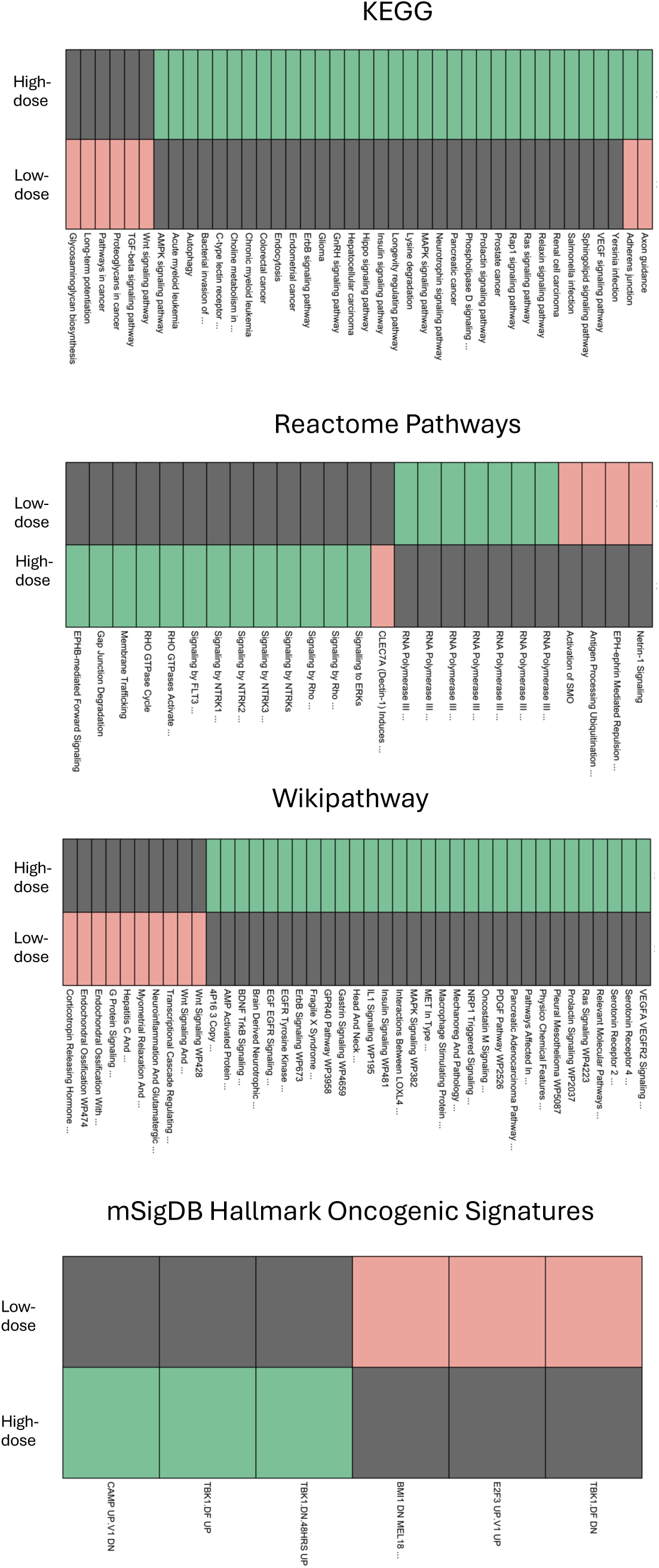
Pathways for significantly enriched DVs from the high- and low-dose arms for each tested data based (KEGG, Wikipathway, Reactome, mSigDB Hallmark Oncogenic signatures).

**Supplementary Tables** (*see supplementary files*):

**Table S1.** High-dose hypomethylated DMR enrichment.

**Table S2.** Pathway analysis using other DMR sets.

**Table S3.** High-dose and low-dose immune panel DEGs.

**Table S4.** DV enrichment results

## Notes

The authors declare no potential conflicts of interest.

### Competing Interest Statement

The authors have declared no competing interest.

### Clinical Trial

NCT02295059

### Clinical Protocols

https://clinicaltrials.gov/study/NCT02295059

### Funding Statement

This study was funded by R01 CA164019 (LDY). We are grateful for the COHCCC Core Facilities supported by NIH P30CA033572, including the Integrative Genomics Core. Additional support was provided by USDA Project 2032-51530-025-00D. The USDA is an equal opportunity employer and provider. The USDA had no influence in the study design or interpretation of the experimental results. The reported findings and their interpretation are the expressed opinions of the authors and do not represent an official position of the USDA.

### Author Declarations

This study was approved by the Institutional Review Boards of The Ohio State University (OSU) and City of Hope (COH).

